# Evaluation of sample collection and transport strategies to enhance yield, accessibility, and biosafety of COVID-19 RT-PCR testing

**DOI:** 10.1101/2021.03.03.21251172

**Authors:** Padmapriya Banada, David Elson, Naranjargal Daivaa, Claire Park, Samuel Desind, Ibsen Montalvan, Robert Kwiatkowski, Soumitesh Chakravorty, David Alland, Yingda L. Xie

## Abstract

Sensitive, accessible, and biosafe sampling methods for COVID-19 reverse-transcriptase polymerase chain reaction (RT-PCR) assays are needed for frequent and widespread testing. We systematically evaluated diagnostic yield across different sample collection and transport workflows, including the incorporation of a viral inactivation buffer. We prospectively collected nasal swabs, oral swabs, and saliva, from 52 COVID-19 RT-PCR-confirmed patients, and nasopharyngeal (NP) swabs from 37 patients. Nasal and oral swabs were placed in both viral transport media (VTM) and eNAT™, a sterilizing transport buffer, prior to testing with the Xpert Xpress SARS-CoV-2 (Xpert) test. The sensitivity of each sampling strategy was compared using a composite positive standard. Overall, swab specimens collected in eNAT showed superior sensitivity compared to swabs in VTM (70% vs 57%, P=0.0022). Direct saliva 90.5%, (95% CI: 82%, 95%), followed by NP swabs in VTM and saliva in eNAT, was significantly more sensitive than nasal swabs in VTM (50%, P<0.001) or eNAT (67.8%, P=0.0012) and oral swabs in VTM (50%, P<0.0001) or eNAT (56%, P<0.0001). Saliva and use of eNAT buffer each increased detection of SARS-CoV-2 with the Xpert test; however, no single sample matrix identified all positive cases.

## INTRODUCTION

Accurate, efficient, and biosafe detection of SARS-CoV-2 in both symptomatic and asymptomatic individuals with active COVID-19 infection is an essential public health strategy for preventing transmission and controlling the COVID-19 pandemic. Although nasopharyngeal (NP) swabs are a preferred specimen type, the invasiveness of this procedure, potential for variable collection quality, need for supervised collection with biohazard risks have hindered the scalability of this testing method ^1-4^. Rather, there has been progress in establishing utility of non-invasive sampling methods that use saliva ^5-7^ or self-collected nasal or oral swabs ^8, 9^ which can enable broader testing of at-risk populations and decrease exposure risk to healthcare workers ^10^. When combined with self-testing, wider implementation of rapid, CLIA-waived COVID-19 assays^11,12^ could dramatically increase public access to tests for SARS-COV-2 infection by expanding testing in at-risk locations including school and the workplace. However, the feasibility of scaling these non-invasive samples for widespread testing outside of carefully controlled environments remains limited by the operational and biosafety challenges.

Our group has recently demonstrated that a guanidine-thiocyanate transport buffer (eNAT™, Copan diagnostics Murrieta, CA) leads to viral inactivation, with at least 5-log reduction in viable SARS-COV-2, and stabilization of viral RNA in a sample^13^. With the premise that eNAT could optimize testing yield while simplifying sample transport and handling, we evaluated and compared the yield of eNAT versus standard transport media, across different non-invasive samples, using the Cepheid Xpert Xpress SARS-COV-2 test (‘Xpert’). Xpert is a FDA- EUA approved rapid, integrated, cartridge-based RT-PCR test that can be run on widely existing GeneXpert instruments used in over 130 countries. To our knowledge, this is the first study to demonstrate the use of a viral inactivating buffer across various non-invasive samples in a point-of-care test to deliver a complete and scalable work flow for biosafe handling and testing of COVID samples.

## MATERIALS AND METHODS

### Study population and sample collection

To collect SARS-CoV-2 positive samples from COVID-19 PCR confirmed participants, we conducted a sub-study to an observational cohort study of COVID-19 patients at University Hospital (UH) affiliated with the Rutgers New Jersey Medical School in Newark, NJ, USA. This study was approved by the Rutgers Institutional Review Board for human subject research (Rutgers IRB # Pro2020001138). Eligible patients included adults (age ≥ 18) who tested SARS-CoV-2 PCR-positive by the in-hospital NP swab PCR tests (most commonly Simplexa COVID-19 Direct EUA (Diasorin Molecular LLC, Cypress, CA)). Patients that could not or did not consent, were pregnant or breastfeeding, prisoners, or who were unable to provide any respiratory specimens were excluded. Trained study personnel collected 1 NP swab (baseline only), 2 oral swabs, 2 nasal swabs, and a saliva sample from all participants who consented to all sample types. A subset of participants being evaluated for hospital outcomes in the parent study continued to be sampled longitudinally by oral swabs, nasal swabs, and saliva every 2-3 days until discharge. All swab types were immediately placed into 3mL of sterile Universal Viral Transport Medium (VTM; Labscoop, Little Rock, AR) whereas a second nasal and oral swab was collected and immediately placed into 3mL of eNAT (COPAN Diagnostics, Murrieta, CA, USA). A thinner nylon tip swab designed for nasopharyngeal (NP) sampling was used to obtain the NP swab (baseline only), and a thicker nylon tip swab designed for oral and nasal samples (Copan diagnostic, Murrieta, CA) was used for these sample types. NP swab collection was performed in accordance with CDC guidelines ^14^; oral swab collection was performed by swabbing both buccal surfaces and tongue with an alternating order of collection for each media; nasal (anterior nares) swab collection was performed by rotating the swab 1 cm inside the nostril for 10-15 seconds, alternating nostrils for each media. Additionally, participants were instructed to self-collect a posterior saliva sample by clearing the back of their throat, then collecting 4 mL of saliva into a marked, empty, sterile wide-mouth cup (though any volume over 0.5 mL was accepted). All specimens were transported at room temperature and stored in a 2-4°C prior to testing, which occurred within 48 hours of sample collection.

### Testing by Xpert Xpress SARS-Cov-2 (‘Xpert’)

NP, nasal, and oral swabs were tested by adding 300µl of the sample (either in VTM or eNAT) directly to the Xpert SARS-Cov-2 test cartridges and the test was run in the GeneXpert system as per the manufacturer’s instructions. The saliva sample was tested using three different methods. First, 300µl of the saliva sample was directly added into the Xpert test cartridge (“saliva direct” sample). Additionally, the same saliva sample was swabbed with two separate swabs (thicker nylon tip swabs) for 10 twirls followed by incubating each swab in the saliva for ∼10-20 seconds (“saliva swab” sample). Each saliva swab sample was then transferred into test tubes containing 3mls of either VTM or 2 ml of eNAT buffer and mixed well. From each of these mixtures, 300µl were added directly to the sample chamber of Xpert cartridges. Saliva samples <300 µl were tested only by swabbing in eNAT and VTM. We also compared saliva samples directly diluted in 1:1, 1:2 and 1:4 ratios of saliva to eNAT. A minimum volume of 700µl of saliva was needed to test all saliva processing methods: ‘saliva direct’, saliva swab in eNAT and all three dilutions. For saliva samples with volumes less than 700µl, we prioritized saliva direct and saliva swab testing. Out of the 44-saliva direct positive samples tested with eNAT ratios, 1:1 dilution was not performed for one saliva sample due to insufficient volume. One each of the sample types had an error either due to pressure aborts (Error 2008) or probe check error (Error 5017) or instrument hardware error (Error 2025) and were not repeat tested. The saliva:eNAT mixtures were then tested using the GeneXpert system by adding 300µl of the mixture to the Xpert SARS-CoV-2 test cartridge. The effect on the assay inhibition, N2 gene cycle threshold (Ct), percent positive rate and cartridge pressure values were evaluated.

### Definitions

We compared samples that were collected contemporaneously (sample comparison set) and applied a composite SARS-CoV-2 positive reference standard, defined as at least one sample type being positive in the sample comparison set. We did not compare sample sets in which no samples were positive, as we reasoned that the PCR-negative samples from these individuals could not be considered false negative due to biologic variability in sampling over time, but they were also not suitable as true negative comparators due to known COVID status of these individuals. To confirm the discordancy that negative comparison sets was not due to difference in the in-hospital versus Xpert PCR tests, we obtained leftover media from positive NP swabs of a random subset of six participants. We then tested this archived sample as validation samples on Xpert. Xpert correctly detected SARS-CoV-2 in all six of these archived samples.

### Statistical analyses

Standard statistical analyses (average, standard deviation, and t-test) and proportion of positive tests by each sampling method were compared by Chi-square, t-test or z-test as appropriate, using Microsoft Excel 365 for Windows, GraphPad Prism version 8 or online software (http://vassarstats.net/propdiff_ind.html). Scatter plots for Ct values showing the mean and SD were included for the positive samples.

## RESULTS

### Participant enrollment and characteristics

Between June 12^th^, 2020 and October 23^rd^, 2020, 70 subjects were enrolled into the study (Figure 1). From these 70 enrollees, a total of 116 sample comparison sets were collected - 70 at baseline and 46 at follow-up time-points. Of note, some participants consented to all sample types except for NP swabs. Of the 116 comparison sets, 84 sample sets from 52 participants were complete with all specimen types and had at least one sample positive (by the composite reference standard) and were thus included in the sample comparison analysis (Fig. 1). Characteristics of the 52 participants in the analysis population (participants with at least one study sample positive for SARS-CoV-2) are shown in Table 1 and characteristics of the 13 participants with all negative samples are shown in Supplementary Table S1. Among the 52 participants in the analysis population, 41 (79%) had symptoms potentially consistent with COVID whereas 11 (21%) of these participants presented to the hospital for non-COVID indications, had no respiratory symptoms, and were incidentally found to be COVID-positive. Average participant age was 55, 37% were female, and the most common comorbidities were hypertension and diabetes.

**Figure 1.**
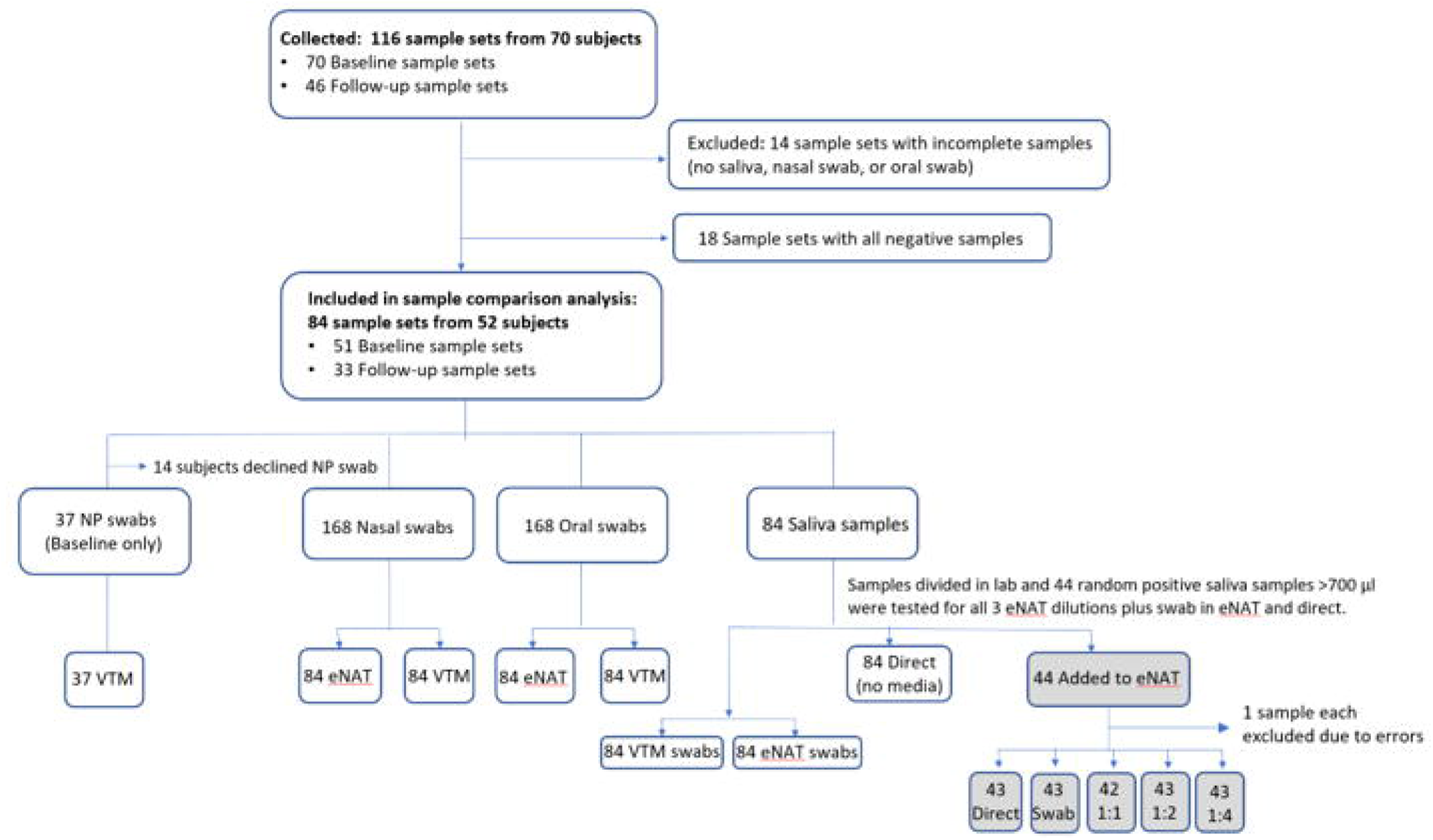
Study flowchart.

**Table 1.** Characteristics of participants in the analysis population (participants with at least one study sample positive for SARS-CoV-2).

|  | Analysis population<br>(N=52) |
| --- | --- |
| Mean Age in years (SD) | 55 (15.1) |
| # of Men (%) | 33 (63%) |
| # of Women (%) | 19 (37%) |
| Ethnicity (%) |  |
| Hispanic | 35 (67%) |
| Black | 15 (29%) |
| White | 2 (4%) |
| Comorbidities |  |
| Hypertension | 27 (52%) |
| Diabetes Mellitus | 16 (31%) |
| Coronary Artery Disease | 7 (13%) |
| Chronic Kidney Disease | 4 (8%) |
| Lung Disease (eg, COPD) | 8 (15%) |
| No chronic disease | 19 (36%) |
| COVID symptoms (%) |  |
| Cough | 33 (64%) |
| Shortness of breath | 32 (62%) |
| Fever | 31 (60%) |
| Diarrhea | 13 (25%) |
| Chest Pain | 10 (19%) |
| No COVID symptoms | 11 (21%) |
| Oxygen Support Required (%) |  |
| None | 20 (38%) |
| Nasal Canula | 29 (56%) |
| Non-Invasive Mechanical Ventilation | 2 (4%) |
| Intubation | 1 (2%) |
| Symptom duration prior to baseline collection<br>Mean (range) | 7 days (1 – 23 days) |
| Days between in-hospital NP swab PCR and<br>baseline collection: mean (range) | 2 days (0 – 10 days) |
| Number of follow-up time-points per participant:<br>mean (range) | 1.5 (0 – 10) |
| Participants with negative NP swab PCR collected<br>in clinical follow-up during hospitalization | 19 (38%) |

On average, the baseline collection took place 2 days after the last positive in-hospital NP swab PCR test for participants in the analysis group, and 3 days for participants with no positive samples. The biologic variability of PCR positivity from samples collected several days apart was evident in the discordancy of longitudinal in-hospital NP swab PCR testing results even when the same test was used. Nineteen (38%) of the 52 participants in the analysis group had at least one subsequent negative in-hospital NP swab PCR test during their hospital admission (Table 1). Additionally, we validated 100% agreement of the in-house test with Xpert (all the original samples were Xpert positive) from the original left-over positive NP swab specimen of 6 participants. These observations support that positive-negative discordancy across time was likely biologic or sampling variability and unlikely due to discordancy between the in-hospital and Cepheid tests, and is consistent with previous comparative performance of Xpert Xpress SARS - CoV-2 with other SARS-CoV-2 RT-PCR platforms ^11, 15, 16^.

### Comparative testing of different respiratory specimens in Xpert Xpress SARS-COV-2

A total of 84 sample comparison sets from 52 patients were included in the sample comparison analysis based on the composite reference, where at least one specimen in the comparison set was positive. Seventeen of these patients had follow-up samples collected on alternative days during their hospital stay. Thus, a total of 84 sets of all specimen types were included in the analysis except for NP swabs, which were only collected from 51 completed baseline collections. Of the 51 completed baseline collections, 14 participants declined NP swab, leaving a total of 37 sample sets that could be analyzed with NP swab.

As shown in Fig. 2A, undiluted saliva added directly to the cartridge (‘direct saliva’) gave the highest detection rate at 90.5% (76/84), followed by NP-VTM (86.5%, 32/37) and saliva in eNAT buffer (84.5%; 71/84), which were significantly higher compared to nasal or oral swabs (P<0.0001). Saliva in VTM (71.4%; 60/84) also performed better than oral swabs in VTM (50%; 42/84) or eNAT (58%; 49/84), as well as nasal swabs in VTM (50%; 42/84) or eNAT (67.8%; 57/84). We further analyzed N2-gene cycle threshold (Ct) values for all positive samples as shown in Fig. 2B. Average N2 gene Ct values were the earliest for NP-VTM (32±5.4) and saliva direct (Ct=34.2±5.8) and most delayed for oral-VTM (37.5±4.9). The Ct range difference was statistically significant between saliva direct and oral-VTM (P<0.0001), oral-eNAT (P=0.0003) and saliva-VTM (P=0.0026). However, there was no significant difference of N2-Ct range for NP-VTM (P=0.28), nasal-VTM (P=0.09), nasal-eNAT (P=0.82) and saliva-eNAT (P=0.26) compared to saliva direct (Fig. 2B). There were three negative NP specimens that were detected in saliva, which we observed to have N2 Ct values of 39.4, 40.3 and 36.1 (Supplementary Fig. 1C), indicating below LOD level viral loads ^17^ possibly contributing to the discrepancy. Only one of the sample sets was positive by NP swab (Ct=35.4) but negative in saliva direct and both saliva swabs (VTM and eNAT). Overall, we found that saliva performed better or equal to NP swabs in detecting COVID positive patients. Similarly, the samples that were negative by other respiratory specimens (nasal or oral swab) but detected by saliva swab in VTM or eNAT had an overall delayed N2-Ct values of >37, indicating better performance in saliva for samples with low viral load (sub-LoD) or lesser variation in saliva collection.

**Figure 2.**
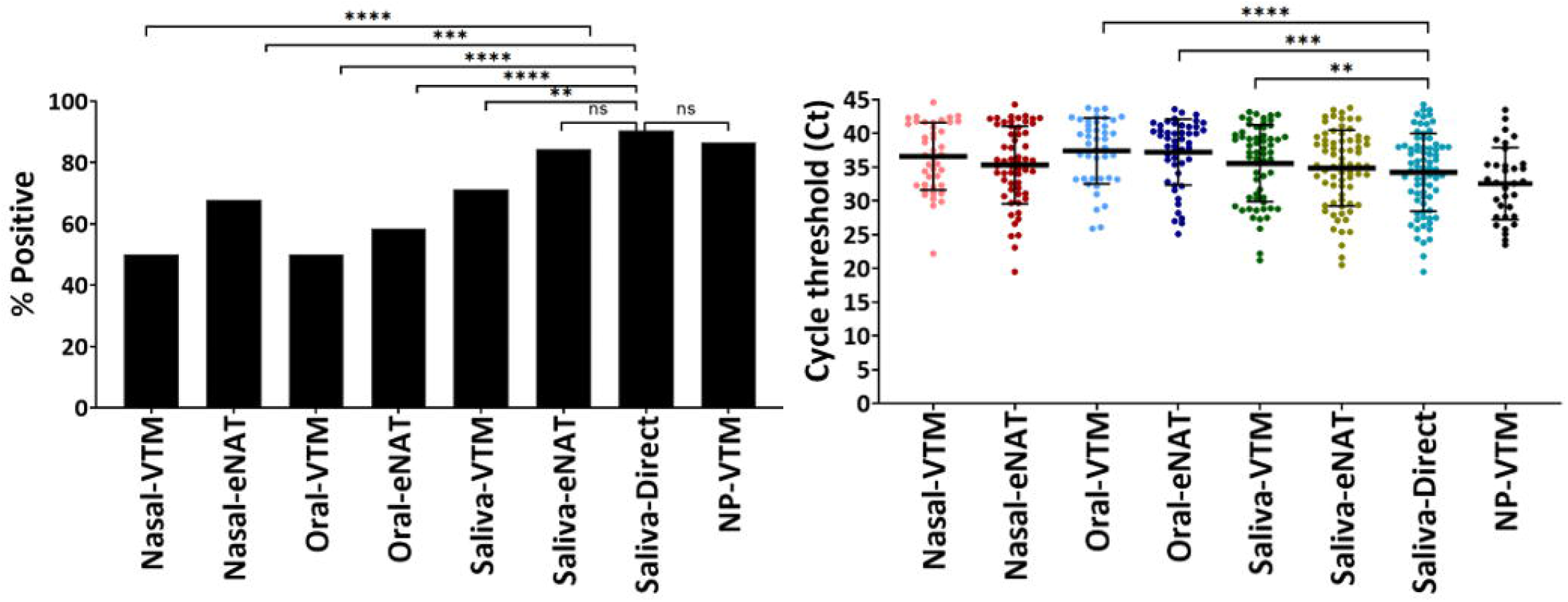
Comparative testing of different respiratory specimens using the Xpert Xpress SARS - COV-2 test. (A) Percent positive rate and (B) N2 gene cycle threshold (Ct) values of samples from all participants with at least one SARS-COV-2 positive sample (N=84 for all samples and N=37 for NP swab). NP=Nasopharyngeal: VTM=Viral transport medium; eNAT= eNAT™ transport media, Copan diagnostics. ns=not statistically different. **** P<0.0001; ***P<0.001, **P=0.02

### Influence of transport media on detection across all sample types

We also evaluated if the composition of different transport media, specifically VTM and eNAT, had any influence on the detection sensitivity. As described, nasal and oral swabs were collected in both VTM and eNAT whereas saliva was collected from patients in an empty sterile cup, then subsequently swabbed and stored in VTM and eNAT. As shown before in Fig. 2A, compared to VTM, eNAT increased the positivity rate by about 20% (40/84 vs 57/84) for nasal swabs (P=0.008), followed by 12% for saliva (60/84 vs 70/84, P=0.065) and 6% for oral swabs (42/84 vs 47/84, P=0.43). When data from all sample types were combined to compare the two media, eNAT offered over 12% advantage (142 vs 174 out of 252 samples) in overall detection rate compared to VTM (P=0.003).

### Optimizing the use of eNAT buffer for saliva

Compared to saliva swabbed into eNAT, direct saliva yielded an overall delayed SPC-Ct values in the Xpert test, indicating possible PCR inhibition and increased (>60 PSI) in-cartridge pressure values (Supplementary Fig. 2B). Saliva diluted into eNAT at a ratio of 1:2 (N=43) yielded the second highest PCR positive rate (97.7%, 42/43) after saliva direct (100%, 43/43) (Fig. 3). Dilutions of 1:1 and 1:4 yielded 95% (40/42) and 93% (40/43) positive rate, respectively. Saliva swabs in eNAT showed the lowest sensitivity at 86% (37/43, P>0.05; Fig. 3A). A sample missed by 1:2 &1:4 dilutions and another by 1:1 & 1:4 dilutions, had delayed N2-Ct values of 44.3 and 41.7 with saliva direct, respectively, indicating the influence of Poisson distribution for viral loads considerably below the limit of detection. Whereas the average N2-Ct values were similar (ca. 33-34) for all saliva conditions tested (P>0.05), the SPC-Ct values were earlier with saliva in eNAT compared to saliva direct (31.25±1.74, P<0.001, Supplementary Fig. 2C), suggesting that PCR inhibition was mitigated by the addition of eNAT to an appreciable extent.

**Figure 3.**
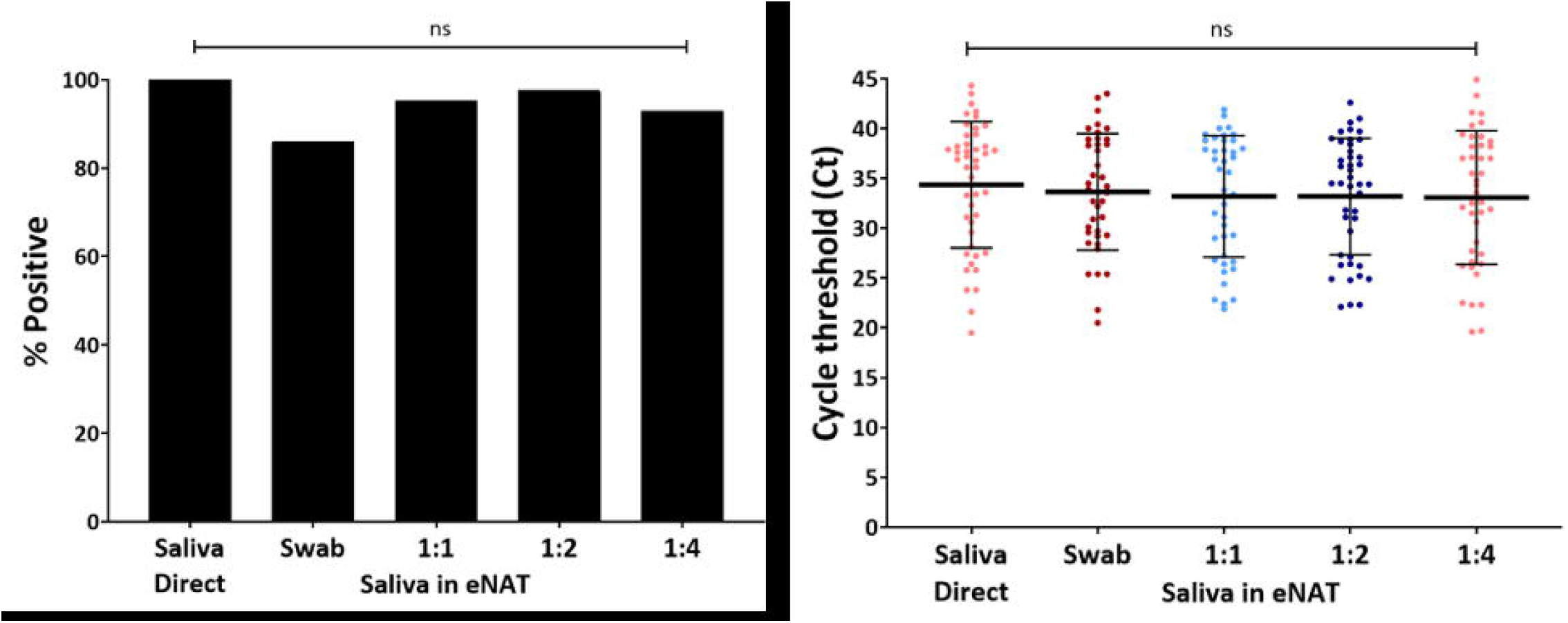
eNAT as a transport media for saliva. Saliva diluted with eNAT at 1:1, 1:2, and 1:4 ratio showing (A) Percent positive rate and (B) N2-Ct values from patient saliva samples tested directly (N=44), as a swab in eNAT (N=44), diluted 1:1 (N=44), 1:2 (N=42), and 1:4 (N=42) in eNAT transport media. ns=not statistically significant.

### Operational characteristics of processing saliva in GeneXpert cartridges

To evaluate saliva processing profiles in the GeneXpert cartridges, we analyzed the sample processing control (SPC) Ct values and in-cartridge pressure values. All respiratory samples collected in either VTM or eNAT did not have any significant difference either with SPC Ct or the max pressure values (Supplementary Fig. 2A and 2B). Saliva direct, the only sample type analyzed as is without dilution, yielded slightly delayed SPC Ct and higher cartridge pressure values with an average of 58±13.48, with one sample aborting the run due to pressure exceeding 100 psi (vs NP-VTM, P<0.0001). However, when the saliva was swabbed and transferred to VTM or eNAT, average pressure values fell to 53.2±6.06 (P<0.0001) and 52.2±5.4 (P<0.0001), respectively. Dilution with eNAT at 1:1, 1:2 and 1:4 ratio reduced the inhibition from saliva direct (P<0.0001) by lowering the average SPC-Ct values by ∼2 ct values (Ct 29.1 in 1:2 vs 31.2 in saliva direct). There was no significant difference in maximum in-cartridge pressure values with saliva dilution in eNAT (P>0.05), except for swab in eNAT (P=0.02). These results suggest that particles or mucus present in direct saliva samples can occasionally interfere with assay function, and that swab testing may be considered when these situations occur.

## DISCUSSION

We found that saliva is an excellent test matrix for the Xpert Xpress SARS-CoV-2 test, providing a sensitivity that is comparable to NP swabs and better than nasal and oral swabs. This finding is consistent with previously published studies using other RT-qPCR modalities ^18-23^. Although a handful of previous studies have looked at saliva tested in the Xpert SARS-CoV-2 ^11, 24, 25^, to our knowledge this study is the first study to comprehensively test multiple non-invasive sampling methods, in the setting of both symptomatic and asymptomatic SARS-COV-2 infection, with and without the use of a sterilizing sample/transport buffer. By applying a composite reference standard for a positive sample, we observed that saliva enhanced the detection of SARS-CoV-2 compared all other sampling types. This finding may be largely attributable to more reliable SARS-CoV-2 yield with saliva sampling as opposed to higher variability of swab sampling, consistent with similar observations from other studies ^1, 5-7^. It is worth noting that noB sample matrix was 100% sensitive compared to the composite reference standard. Discordancy between sample matrices was most pronounced in samples that had a delayed cycle threshold indicating low viral load. This suggests that testing with multiple samples and perhaps multiple sample types when clinical suspicion is high may provide the highest sensitivity and negative predictive value for SARS-CoV-2.

We additionally found that eNAT, a buffer we have previously determined to be effective at inactivating SARS-CoV-2 in-vitro (Banik et al., submitted), increased the test positivity rates across all sample types compared to VTM (P=0.0032), with a saliva to eNAT ratio of 1:2 being optimal in our sample set. We also found that adding eNAT to saliva possibly mitigates the PCR interference from saliva with lower pressure values and recovery of otherwise delayed SPC Ct values seen with direct saliva. These findings suggest that the application of eNAT as a sample buffer may be advantageous not only in safe handling and transport, but also in improving yield and processing capability on the Cepheid system.

There were several limitations in this study. First, there were less contemporaneous NP swabs collected with saliva, thereby reducing the number of direct comparisons between these two sample types, although they were found to be comparable. An underlying reason for this – participants declining NP swab collection due to its discomfort – also demonstrates the real-world limitations that would be magnified with larger scale testing such as in schools or the workplace. Secondly, we added eNAT to saliva in the laboratory, whereas the benefit of eNAT would be to sterilize samples immediately after collection and before transport and test set up. However, this allowed us to evaluate the combination of eNAT and saliva under different conditions and inform optimal design of kits to add eNAT immediately to saliva upon collection. Finally, our participants were patients who had either been admitted to the hospital or seen in the emergency department. This population may not be generalizable to ambulatory individuals who would benefit the most from self-collection. However, we captured a diverse patient group in our cohort including those who were never admitted, as well as patients who were detected by universal screening but reported no COVID symptoms.

Altogether, our findings support the use of saliva and eNAT sterilizing buffer to enhance effective, safe, and accessible COVID testing and screening in the many health care systems worldwide already using GeneXpert instruments.

## Data Availability

The data that support the findings of this study are available from the corresponding author, YLX and DA, upon reasonable request.

## Acknowledgements

We thank Dr. Jason H. Yang (Rutgers New Jersey Medical School) for supporting DE’s and CP’s role in the studies, Cepheid for in-kind donation of cartridges, and Copan for donation of eNAT media and swabs. This study was partially funded by the National Institute of Allergy and Infectious Diseases of the National Institutes of Health under award number R01 AI131617 and Rutgers University Institutional Support.

