## Supplemental materials for "Evaluation of sample collection and transport strategies to enhance yield, accessibility, and biosafety of COVID-19 RT-PCR testing"

| | Individuals with all negative samples (N=13) |
|---|---|
| **Mean Age in years (SD)** | 57 (15) |
| **# of Men (%)**<br>**# of Women (%)** | 8 (61%)<br>5 (39%) |
| **Ethnicity (%)**<br>Hispanic<br>Black<br>White | <br>5 (39%)<br>6 (46%)<br>2 (15%) |
| **Comorbidities**<br>Hypertension<br>Diabetes Mellitus<br>Coronary Artery Disease<br>Chronic Kidney Disease<br>Lung Disease (eg, COPD)<br><br>No chronic disease | <br>5 (39%)<br>2 (15%)<br>1 (8%)<br>3 (23%)<br>2 (15%)<br><br>5 (38%) |
| **COVID symptoms (%)**<br>Cough<br>Shortness of breath<br>Fever<br>Diarrhea<br>Chest Pain<br><br>No COVID symptoms | <br>3 (23%)<br>4 (31%)<br>4 (31%)<br>2 (15%)<br>0 (0%)<br><br>8 (62%) |
| **Oxygen Support Required (%)**<br>None<br>Nasal Canula<br>Mechanical Ventilation | <br>9 (69%)<br>0 (0%)<br>4 (31%)<br>0 (0%) |
| Symptom duration prior to baseline collection<br>**Mean (range)** | 12 days<br>(4 – 28 days) |
| Days between in-hospital NP swab PCR and baseline collection: **mean (range)** | 3 days<br>(1 – 5 days) |
| Number of follow-up time-points per participant: **mean (range)** | 0 |
| Participants with negative NP swab PCR collected in clinical follow-up during hospitalization | 6 (50%) |

Table S1. Characteristics of participants with all negative samples.

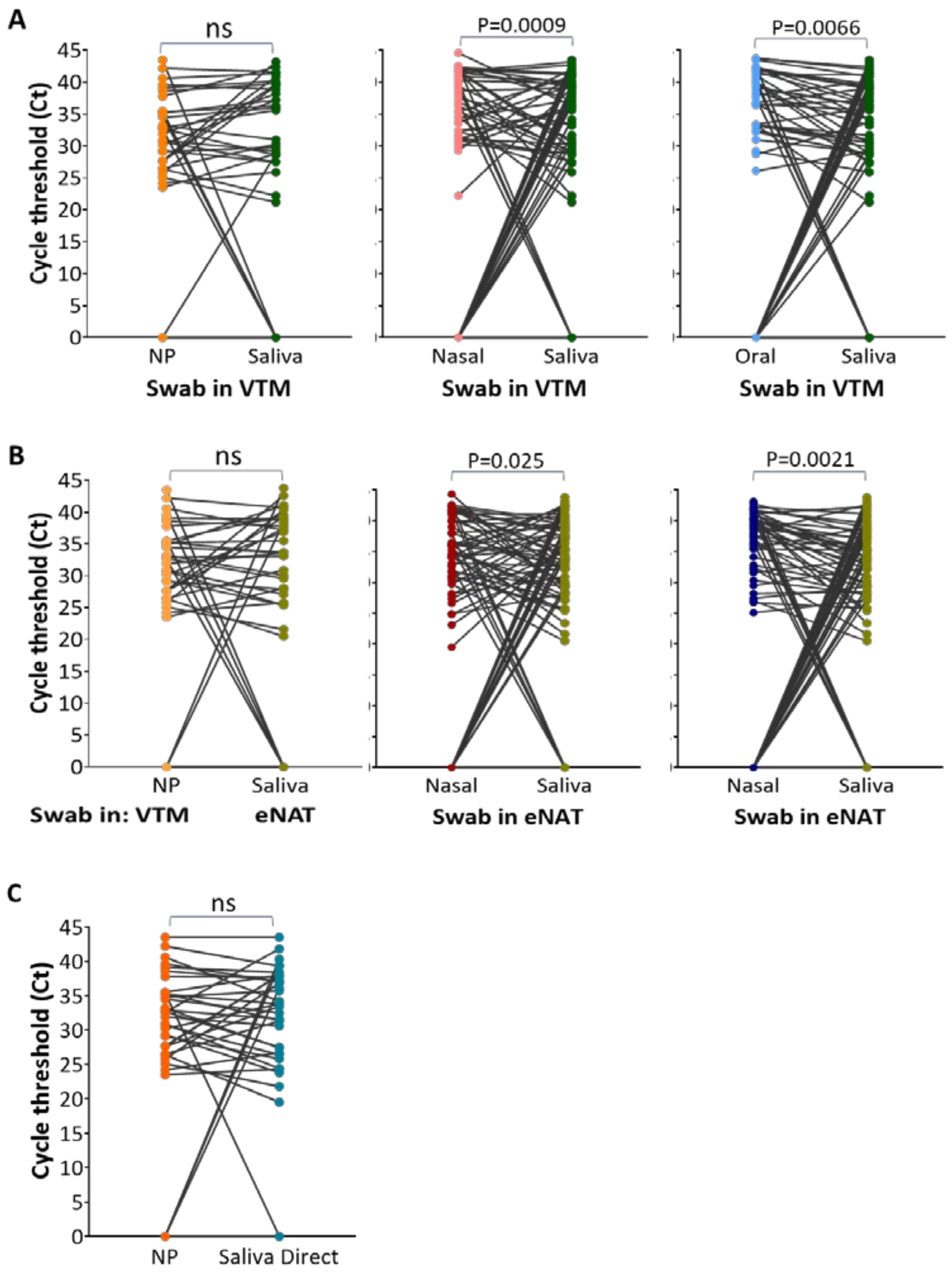


Fig. S2. Paired sample sets showing Ct values comparing each respiratory sample collected in (A) VTM, N=84 and (B) eNAT, N=84. (C) Comparing nasopharyngeal specimens with direct saliva (N=37). The lines indicate samples from the same patient collected at the same time point. ns=not significant.

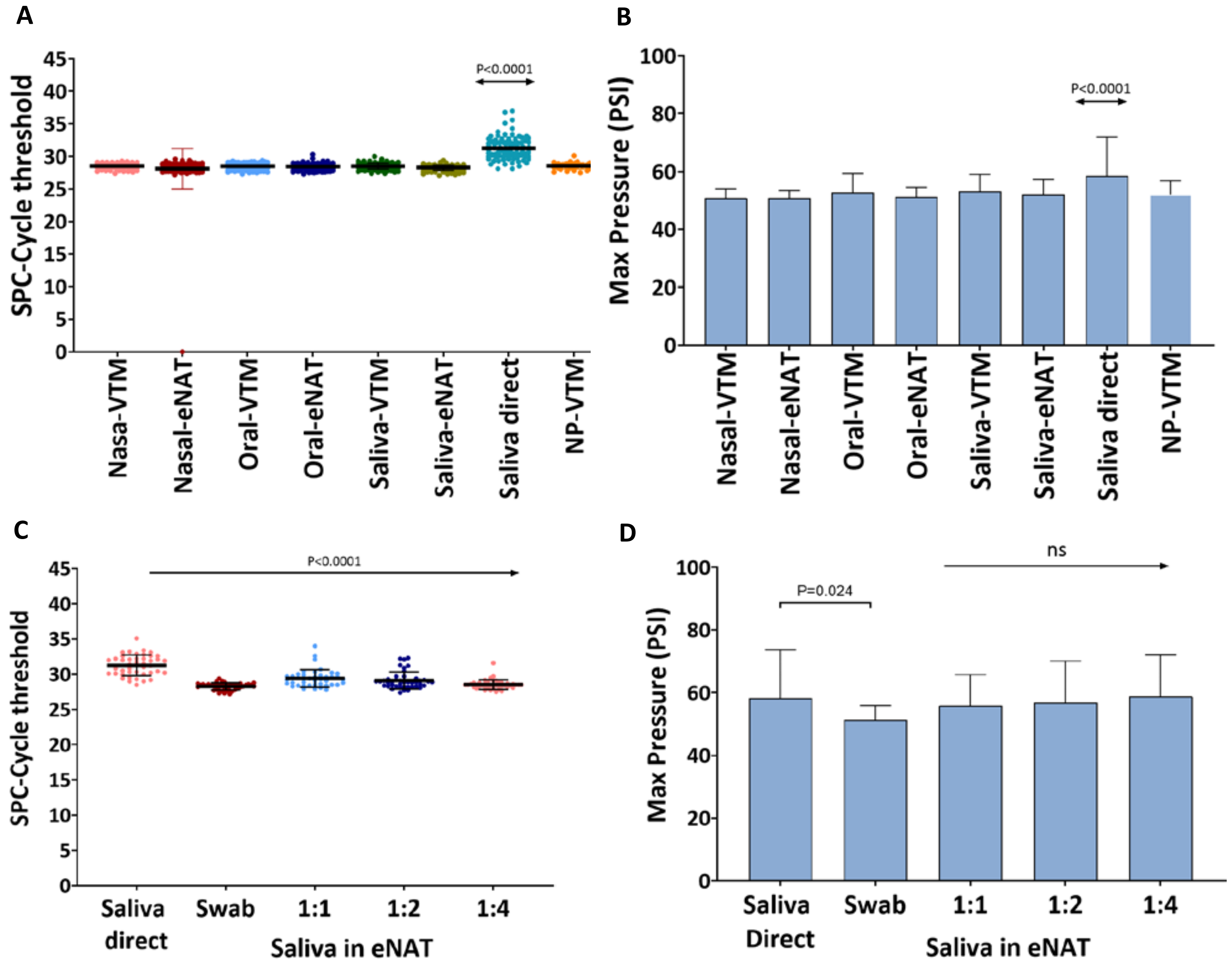


Fig. S3. (A and C) Internal control (SPC) cycle threshold (Ct) values for all respiratory specimens (A, N=84) and for selected saliva samples diluted in eNAT (C, N=44); (B and D) In-cartridge maximum pressure values (PSI) for different respiratory specimens (B, N=84) and for selected saliva samples diluted in eNAT (D, N=44). Each sample tested is represented by points (♦) on the scatter plot and the lines show the mean and standard deviation values. ns= not statistically significant.
